# Increase in preterm stillbirths and reduction in iatrogenic preterm births for fetal compromise: a multi-centre cohort study of COVID-19 lockdown effects in Melbourne, Australia

**DOI:** 10.1101/2021.10.04.21264500

**Authors:** Lisa Hui, Melvin Barrientos Marzan, Stephanie Potenza, Daniel L. Rolnik, Natasha Pritchard, Joanne M. Said, Kirsten R Palmer, Clare L. Whitehead, Penelope M. Sheehan, Jolyon Ford, Ben W. Mol, Susan P. Walker

## Abstract

**Objectives:** The COVID-19 pandemic has been associated with a worsening of perinatal outcomes in many settings due to the combined impacts of maternal COVID-19 disease, disruptions to maternity care, and overloaded health systems. In 2020, Melbourne endured a unique natural experiment where strict lockdown conditions were accompanied by very low COVID-19 case numbers and the maintenance of health service capacity. The aim of this study was to compare stillbirth and preterm birth rates in women who were exposed or unexposed to lockdown restrictions during pregnancy.

**Design:** Retrospective multi-centre cohort study of perinatal outcomes before and during COVID-19 lockdown

**Setting:** Birth outcomes from all 12 public maternity hospitals in metropolitan Melbourne

**Inclusion criteria:** Singleton births without congenital anomalies from 24 weeks’ gestation. The lockdown-exposed cohort were those women for whom weeks 20- 40 of gestation would have occurred during the lockdown period of 23 March 2020 to 14 March 2021. The control cohort comprised all pregnancies in the corresponding periods one and two years prior to the exposed cohort.

**Main outcome measures:** Odds of stillbirth, preterm birth (PTB), birth weight < 3^rd^ centile, and iatrogenic PTB for fetal compromise, adjusting for multiple covariates.

**Results:** There were 24,017 births in the exposed and 50,017 births in the control group. There was a significantly higher risk of preterm, but not term, stillbirth in the exposed group compared with the control group (0.26% vs 0.18%, aOR 1.49, 95%CI 1.08 to 2.05, P = 0.015). There was also a significant reduction in preterm birth < 37 weeks (5.93% vs 6.23%, aOR 0.93, 95%CI 0.87 to 0.99, P=0.03), largely mediated by a reduction in iatrogenic PTB for live births (3.01% vs 3.27%, aOR 0.89, 95%CI 0.81 to 0.98, P = 0.015), including iatrogenic PTB for suspected fetal compromise (1.25% vs 1.51%, aOR 0.79, 95%CI 0.69 to 0.91, P= 0.001). There was no significant difference in the spontaneous PTB rate between the exposed and control groups (2.69% vs 2.82%, aOR 0.94, 95%CI 0.86 to 0.1.03, P=0.25).

**Conclusions:** Lockdown restrictions in a high-income setting, in the absence of high rates of COVID-19 disease, were associated with a significant increase in preterm stillbirths, and a significant reduction in iatrogenic PTB for suspected fetal compromise.

**Trial registration:** This study was registered as an observational study with the Australian and New Zealand Clinical Trials Registry (ACTRN12620000878976).

## Introduction

The COVID-19 pandemic has disrupted the delivery of maternity care, with a recent systematic review concluding that global maternal and fetal outcomes had worsened during the COVID-19 pandemic.^1^ Some studies have reported increases in stillbirth and reductions in preterm birth,^2-8^ while others reported no changes.^9-13^ These differences are likely due to multiple factors, including differences in study methodology,^14^ resource setting, severity of lockdown restrictions, and COVID-19 caseload.^1^

The city of Melbourne, Australia, which has approximately 4,000 births per month, experienced a prolonged period of fluctuating lockdown restrictions commencing on 23 March 2020 through to 14 March 2021 (Figure 1). The strictest period of lockdown in mid 2020 restricted leaving the house for reasons other than approved essential work, caring for dependents, obtaining medical care or essential food/services. Individuals were allowed one hour outside the home for exercise per day within a 5km radius, with a prohibition on all gatherings of more than two people, and a curfew from 8pm to 5am.^15^

**Figure 1.**
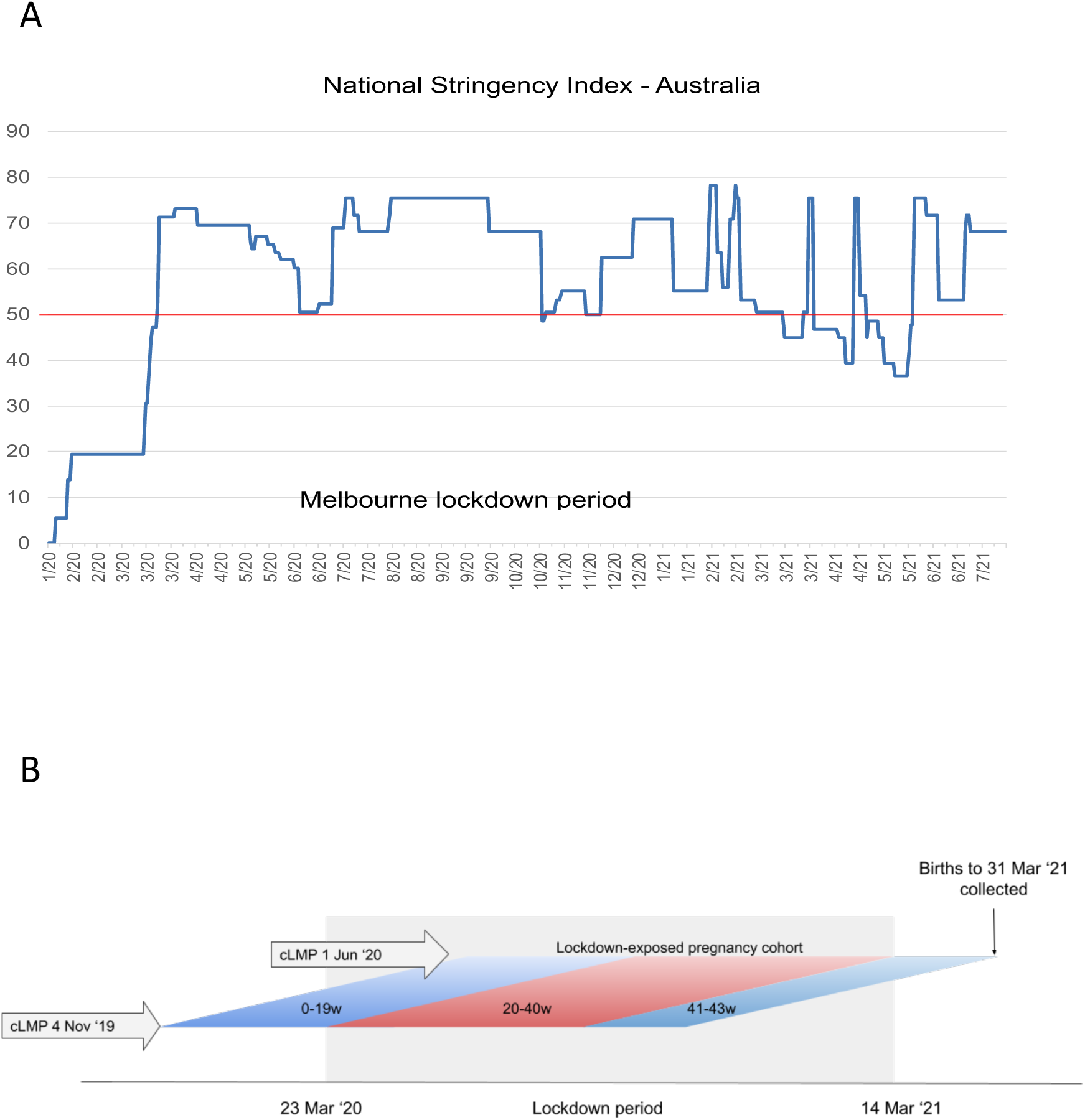
National stringency index and cohort timeline. A. National stringency index for Australia by year/month from the Oxford Government Response Tracker. B. Lockdown-exposed cohort timeline

Numerous modifications to pregnancy care were concurrently adopted to mitigate the anticipated strain on health services and reduce infection risks. These measures included rapid transition to telehealth,^16^ hospital visitor restrictions, increasing the interval between in-person visits, reducing face-to face appointment time, changes to gestational diabetes screening and ultrasound surveillance of fetal growth.^17^ Melbourne experienced relatively few maternal COVID-19 infections (less than 100 in 2020), and no associated maternal or perinatal deaths.^18^ This meant that metropolitan Melbourne experienced a unique set of circumstances not seen in other high-income countries: a prolonged period of significant social restrictions and major changes to antenatal care, without an associated high burden of COVID-19 infections.

In mid 2020, all 12 Melbourne public maternity hospitals (7 health services) formed the *Collaborative Maternity and Newborn Dashboard (CoMaND) for the COVID-19 pandemic* project to internally monitor the effect of the pandemic on clinical quality indicators. Perinatal data collected for CoMaND were used here to analyse the impact of lockdown on preterm birth, stillbirth and utilisation of maternity services.

## Methods

### Ethical approval

This study was given ethical approval from the Human Research Ethics Committees of Austin Health (Ref. HREC/64722/Austin-2020) and Mercy Health (Ref. 2020-031) and is registered as an observational study with the Australian and New Zealand Clinical Trials Registry (ACTRN12620000878976). There was no patient or public involvement in the design of this study.

### Study population

We obtained data from all births ≥ 20 weeks in all 12 public maternity services in Melbourne from 1^st^ January 2018 to 31^st^ March 2021. An estimated 44,000 women give birth in the participating hospitals each year, representing approximately three quarters of all hospital births in Melbourne. Births in exclusively private hospitals and planned home births outside of publicly-funded homebirth programs were not captured. However women planning a private hospital or home birth would typically be transferred to a Level 6 public hospital if they were at risk of preterm birth <31 weeks’ or required tertiary maternal fetal medicine services.

### Exclusions

Infants with congenital anomalies, terminations of pregnancy (TOP), non-Victorian residents, and multiple pregnancies were excluded. We excluded births < 24 weeks’ gestation as TOP can be provided on maternal request up to this gestation, and management of preterm birth and preterm prelabour rupture of membranes < 24 weeks’ is variable and subject to parental discretion.

### Definition of lockdown exposed cohort and non-exposed controls

We defined the lockdown exposure period as 23 March 2020 to 14 March 2021, as this was a continuous period where the national stringency index (NSI) was ≥ 50 on the Oxford COVID-19 Government Response Tracker scale for at least five days per week (Figure 1b).^19^ The NSI threshold of 50 was used in accordance with the definition of lockdown in the International Perinatal Outcomes in the Pandemic study.^20^ Melbourne had the biggest COVID-19 caseload nationally throughout 2020 and therefore drove the Australian government stringency index in 2020.

To maintain anonymity of individual records, hospital data managers converted the actual infant dates of birth into the ordinal calendar week of birth (ie 1 to 52 for each calendar year). For descriptive purposes, weeks are described by the Monday date. We used the calculated week of last menstrual period, rather than week of birth, to define the lockdown-exposed cohort to ensure that outcomes such as preterm birth would be measured using denominator births with a similar timing of lockdown exposure. We subtracted the infants gestational age at birth in completed weeks from the week of birth to obtain the week of calculated LMP (cLMP).

Using this cLMP, we defined a ‘lockdown-exposed’ cohort comprising women for whom weeks 20-40 of gestation would have occurred during the lockdown period. This included women whose cLMP occurred during the 31 weeks from 4^th^ November 2019 to 1^st^ June 2020 inclusive (Figure 1b). To control for possible seasonality, the control group comprised women who had their cLMP during the corresponding calendar weeks commencing one and two years prior to the start of the exposed cohort (births with cLMPs in weeks commencing 5 November 2018 to 3 June 2019 and 6 November 2017 to 4 June 2018). These were assessed as a combined control group.

### Outcome measures

We calculated all outcomes using both denominators of ‘all births’ (live and stillbirths) and ‘live births’, with the exception of stillbirth rate (all births).

### Primary outcomes

1. Total stillbirth rate; stratified by gestational age.
2. Total PTB (< 37 weeks) rate.

### Secondary outcomes

1. PTB < 37weeks: spontaneous and iatrogenic. An iatrogenic birth was defined as any birth without spontaneous onset of labour (either induced labour or no labour).
2. PTB < 32 weeks: total, iatrogenic and spontaneous
3. PTB < 28 weeks: total, iatrogenic and spontaneous
4. Fetal growth restriction (FGR): total FGR, FGR at birth ≥37 weeks, < 37 weeks, < 32 weeks, < 28 weeks. FGR was defined as birthweight < 3^rd^ centile using local population sex-specific birth weight charts.^21^
5. Iatrogenic births for fetal compromise: ≥37 weeks, <37 weeks, <32 weeks, <28 weeks. Indications for induction of labour and cesarean sections (CS) were coded according to the Australian Institute of Health and Welfare Metadata Online Registry, which defines fetal compromise as “suspected or actual fetal compromise, and intrauterine growth restriction”.^22^ Any documentation of suspected fetal growth restriction, antepartum abnormal cardiotocography, “fetal distress” (without labour), reduced fetal movements, oligohydramnios, abnormal umbilical artery Dopplers, or placental insufficiency were included in this classification.
6. Apgar score < 7 at five minutes
7. Special care nursery (SCN) admission
8. Neonatal intensive care unit (NICU) admission
9. First antenatal visit ≤ 12 weeks gestation. This refers to the first planned visit to a midwife or doctor during pregnancy, whether community- or hospital-based.
10. Born before arrival (BBA). This refers to the rate of planned hospital births that occur before arrival, including unplanned births at home, in transit, or other locations.

### Statistical analysis

No sample size calculation was performed as this was a cohort defined by lockdown duration. However, based on prior statewide adjusted stillbirth rates, a total sample size of 64,734 with a 2 to 1 ratio of control to exposed births would allow us to detect a 33% increase in stillbirth from 0.6% to 0.8% with 80% power, at an alpha level of 5%. Analyses of secondary outcomes were considered exploratory and no adjustments for multiple comparisons were made. Perinatal outcomes were summarised as proportion of all births (live births and stillbirths) and live births. Statistical significance was tested with the t-test or chi-square test as appropriate. We performed multivariate logistic regression analysis to obtain the odds ratio (OR) of primary and secondary outcomes in the lockdown-exposed vs non exposed cohorts. We adjusted for the following maternal covariates: maternal age, body mass index (BMI) at first antenatal visit, region of birth, need for interpreter (proxy indicator for primary language and categorised as yes or no), parity, socioeconomic status (assigned by maternal postcode) and smoking in pregnancy status. Adjustments for covariates were done based on the results from backward stepwise regression at alpha <0.15. Records with missing data were excluded from all univariate and multivariable analysis. Statistical analyses were conducted using Stata 17 (StataCorp. 2021. Stata Statistical Software: Release 17. College Station, TX: StataCorp LLC), and two-sided p-values below 0.05 were considered statistically significant. As maternal BMI and smoking status were potentially on a causal pathway between lockdown restrictions and perinatal outcomes, we performed a sensitivity analysis without adjustment for these covariates.

We used Cox regression to derive the hazard ratio of stillbirths and PTB <37 weeks, and Kaplan-Meier curves to plot the cumulative hazard of the outcomes of interest. We tested the proportionality of control and exposed cohorts using tests of non-zero slope. Exponentiated log-ORs of the primary outcomes for each hospital were plotted in forest plots using meta-analysis of aggregated ORs per hospital. We used a fixed effect model to derive the pooled exponentiated effect measures (OR) since all the results are obtained on the same dataset and the heterogeneity across all outcomes was small (0 for all outcomes except overall PTB with I^2^<30).

### Births included in run chart analysis

To examine temporal patterns in outcomes during lockdown conditions, we also generated run charts by week of cLMP. Only births with cLMP in the weeks from 14^th^ August 2017 to 22^nd^ June 2020 were included in the run charts, comprising infants for whom weeks 20 to 43 of gestation occurred within the data collection period of 1^st^ Jan 2018 to 31 March 2021. Pre-pandemic median rates for each outcome were calculated for the non-exposed cohort (cLMP prior to 4^th^ November 2019). A significant shift in a run chart was defined as six or more consecutive weeks all above or below the pre-pandemic median according to established methods for detecting non-random signals in health care.^23^

## Results

There were 147,367 births in participating hospitals during the period 1^st^ January 2018 to 31 March 2021. After classifying all births by week of cLMP, we excluded 16,394 births with cLMP outside the defined period for the run chart analysis. After further exclusions for congenital anomalies, non-Victorian residents, multiple pregnancies and births < 24 weeks, there were 118,705 births remaining for the run chart analysis by week of cLMP (Figure 2).

**Figure 2.**
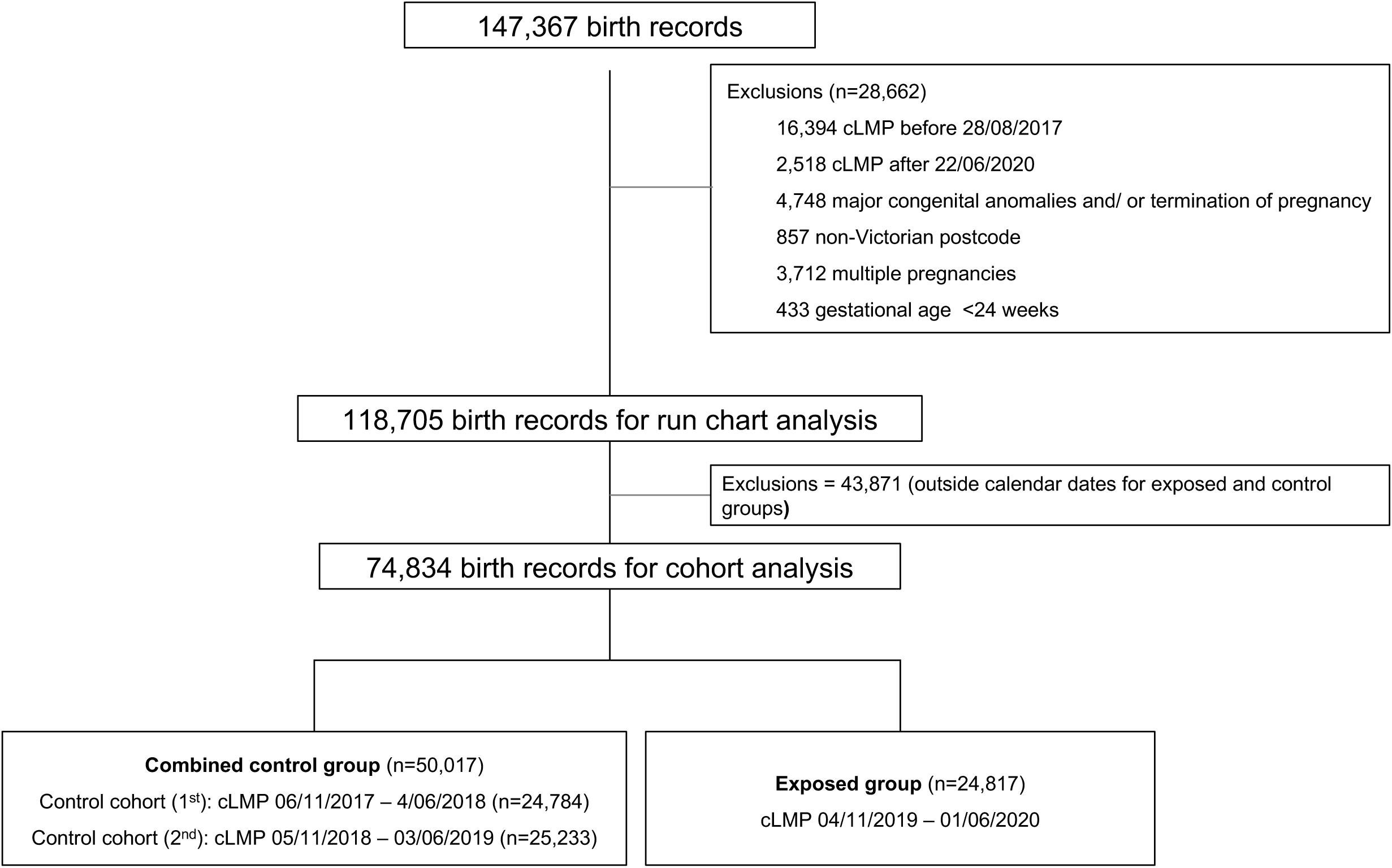
Study flowchart. cLMP, calculated last menstrual period

### Cohort analysis – lockdown exposed vs non exposed pregnancies

The lockdown-exposed cohort contained 24,817 births and the control cohort contained a combined total of 50,017 births (Figure 2). The characteristics of exposed and control groups are shown in Table 1. The lockdown-exposed group differed significantly from the control group in terms of age, socioeconomic class distribution, maternal region of birth, and maternity service level, though the absolute differences were small. Details of the bivariate analysis are available in supplemental file 1. The outcomes are presented using all births denominators (live and stillbirths) in Table 2 and live birth denominators in Table 3. The results all remained robust in the sensitivity analysis excluding covariates of smoking and BMI.

**Table 1.**
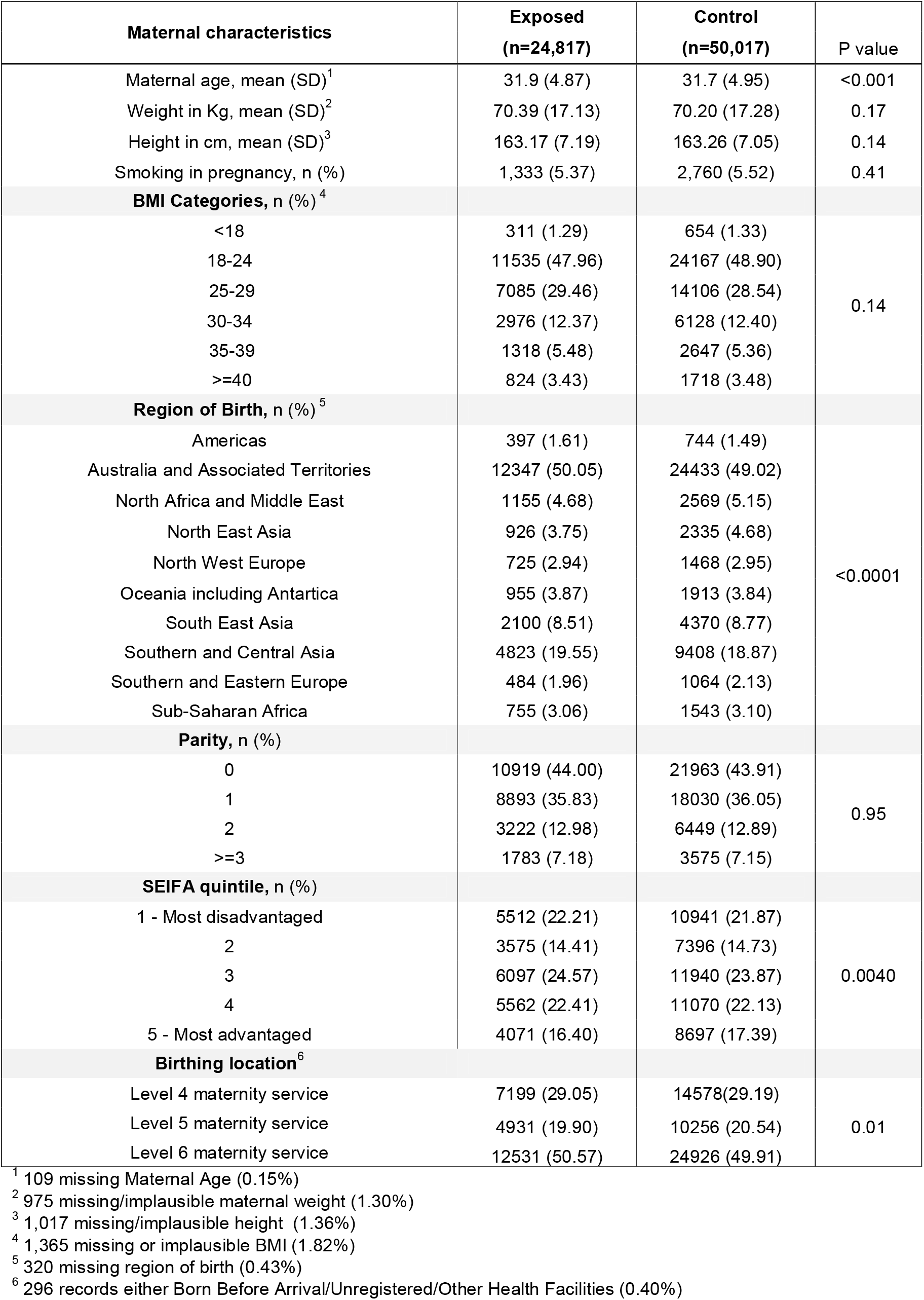
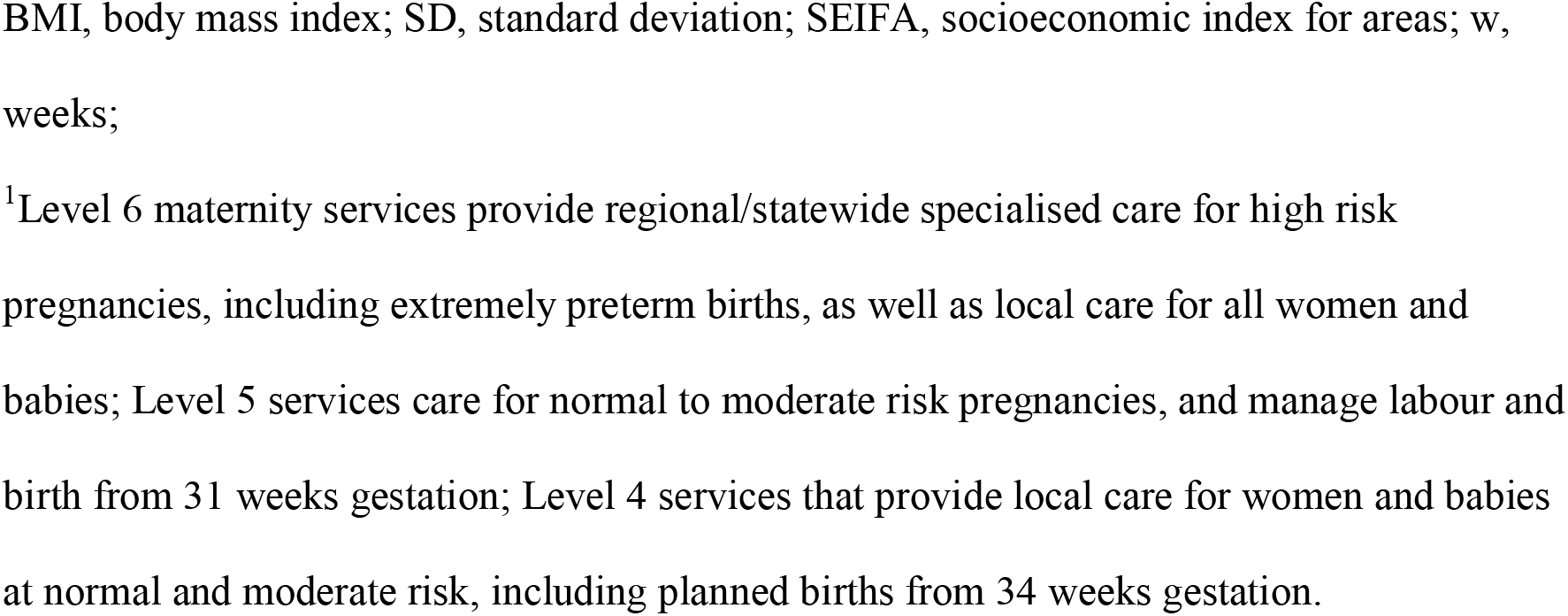
Maternal characteristics among control and exposed cohorts (all births)

**Table 2.**
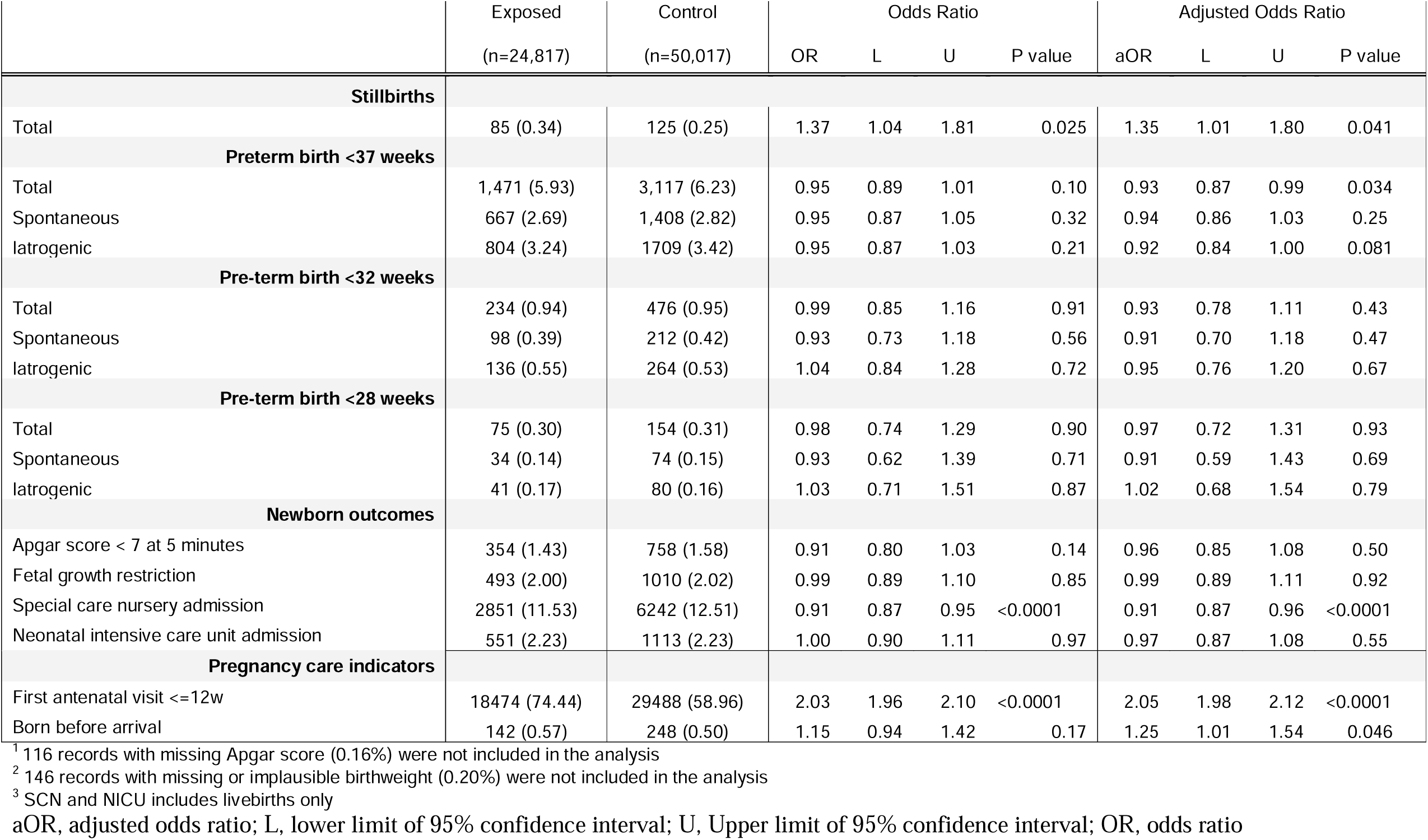
Primary and secondary outcomes for all births (live births and stillbirths)

**Table 3.**
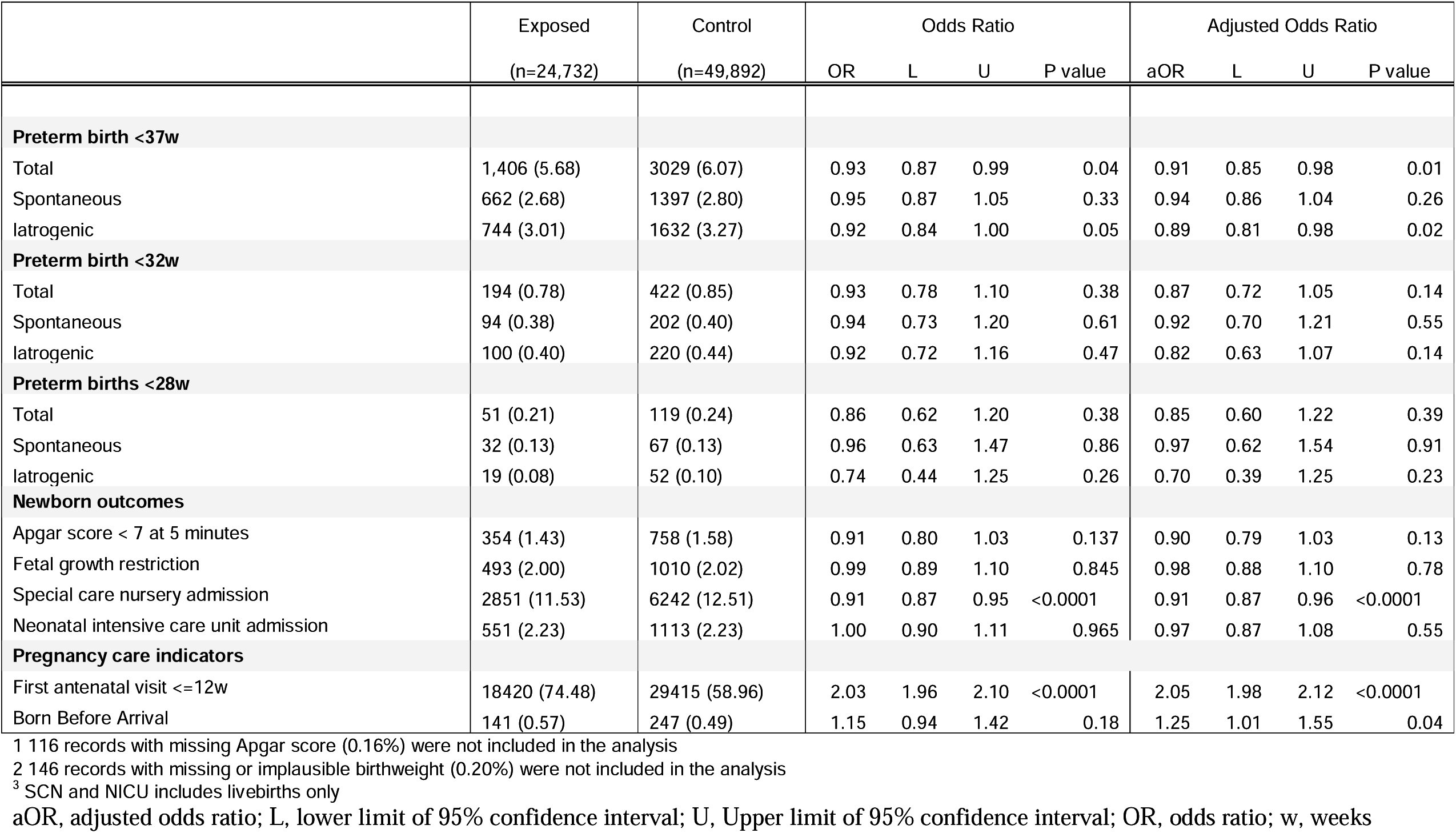
Primary and secondary outcomes for live births.

|  | Exposed<br>(n=24,732) | Control<br>(n=49,892) | Odds Ratio |  |  |  | Adjusted Odds Ratio |  |  |  |
| --- | --- | --- | --- | --- | --- | --- | --- | --- | --- | --- |
|  |  |  | OR | L | U | P value | aOR | L | U | P value |
| **Preterm birth <37w** |  |  |  |  |  |  |  |  |  |  |
| Total | 1,406 (5.68) | 3029 (6.07) | 0.93 | 0.87 | 0.99 | 0.04 | 0.91 | 0.85 | 0.98 | 0.01 |
| Spontaneous | 662 (2.68) | 1397 (2.80) | 0.95 | 0.87 | 1.05 | 0.33 | 0.94 | 0.86 | 1.04 | 0.26 |
| Iatrogenic | 744 (3.01) | 1632 (3.27) | 0.92 | 0.84 | 1.00 | 0.05 | 0.89 | 0.81 | 0.98 | 0.02 |
| **Preterm birth <32w** |  |  |  |  |  |  |  |  |  |  |
| Total | 194 (0.78) | 422 (0.85) | 0.93 | 0.78 | 1.10 | 0.38 | 0.87 | 0.72 | 1.05 | 0.14 |
| Spontaneous | 94 (0.38) | 202 (0.40) | 0.94 | 0.73 | 1.20 | 0.61 | 0.92 | 0.70 | 1.21 | 0.55 |
| Iatrogenic | 100 (0.40) | 220 (0.44) | 0.92 | 0.72 | 1.16 | 0.47 | 0.82 | 0.63 | 1.07 | 0.14 |
| **Preterm births <28w** |  |  |  |  |  |  |  |  |  |  |
| Total | 51 (0.21) | 119 (0.24) | 0.86 | 0.62 | 1.20 | 0.38 | 0.85 | 0.60 | 1.22 | 0.39 |
| Spontaneous | 32 (0.13) | 67 (0.13) | 0.96 | 0.63 | 1.47 | 0.86 | 0.97 | 0.62 | 1.54 | 0.91 |
| Iatrogenic | 19 (0.08) | 52 (0.10) | 0.74 | 0.44 | 1.25 | 0.26 | 0.70 | 0.39 | 1.25 | 0.23 |
| **Newborn outcomes** |  |  |  |  |  |  |  |  |  |  |
| Apgar score < 7 at 5 minutes | 354 (1.43) | 758 (1.58) | 0.91 | 0.80 | 1.03 | 0.137 | 0.90 | 0.79 | 1.03 | 0.13 |
| Fetal growth restriction | 493 (2.00) | 1010 (2.02) | 0.99 | 0.89 | 1.10 | 0.845 | 0.98 | 0.88 | 1.10 | 0.78 |
| Special care nursery admission | 2851 (11.53) | 6242 (12.51) | 0.91 | 0.87 | 0.95 | <0.0001 | 0.91 | 0.87 | 0.96 | <0.0001 |
| Neonatal intensive care unit admission | 551 (2.23) | 1113 (2.23) | 1.00 | 0.90 | 1.11 | 0.965 | 0.97 | 0.87 | 1.08 | 0.55 |
| **Pregnancy care indicators** |  |  |  |  |  |  |  |  |  |  |
| First antenatal visit ≤12w | 18420 (74.48) | 29415 (58.96) | 2.03 | 1.96 | 2.10 | <0.0001 | 2.05 | 1.98 | 2.12 | <0.0001 |
| Born Before Arrival | 141 (0.57) | 247 (0.49) | 1.15 | 0.94 | 1.42 | 0.18 | 1.25 | 1.01 | 1.55 | 0.04 |

<sup>1</sup> 116 records with missing Apgar score (0.16%) were not included in the analysis
<sup>2</sup> 146 records with missing or implausible birthweight (0.20%) were not included in the analysis
<sup>3</sup> SCN and NICU includes livebirths only
aOR, adjusted odds ratio; L, lower limit of 95% confidence interval; U, Upper limit of 95% confidence interval; OR, odds ratio; w, weeks

### Primary outcomes

#### Stillbirths

There was a significantly higher rate of stillbirth in the lockdown-exposed group compared with the control group (0.34% vs 0.25%, adjusted odds ratio (aOR) 1.35, 95%CI 1.01-1.80, P=0.04 (Table 2). When stratified by gestational age, only preterm stillbirths < 37 weeks remained significantly increased in the exposed group (0.26% vs 0.18%, (aOR 1.49, 95%CI 1.08-2.05, P 0.015)(Table 4). The vast majority of fetal deaths were diagnosed prior to the onset of labour; and there was no significant difference in the rates of stillbirths during labour between the exposed and control groups (Table 4). The hazard curve showed a significant gap in the trajectories of lockdown-exposed and control groups in terms of cumulative hazard of stillbirth by gestational age (Figure 3a).

**Table 4.** Stillbirths by timing and gestational age.

| Timing of fetal death | Exposed<br>n = 85 | Control<br>n=125 | Odds Ratio |  |  |  | Adjusted Odds Ratio |  |  |  |
| --- | --- | --- | --- | --- | --- | --- | --- | --- | --- | --- |
|  |  |  | OR | L | U | P<br>value | aOR | L | U | v |
| Before labour | 76 (89.41) | 104 (83.20) | 1.47 | 1.1 | 1.98 | 0.010 | 1.47 | 1.09 | 1.98 | 0.011 |
| During labour | 2 (2.35) | 4 (3.20) | 1.01 | 0.18 | 5.5 | 0.99 | 0.99 | 0.18 | 5.42 | 0.99 |
| Unknown | 7 (8.24) | 17 (13.60) | 0.83 | 0.34 | 2 | 0.68 | 0.83 | 0.34 | 2.01 | 0.68 |
| Gestational age at birth | Exposed<br>n = 24,817 | Control<br>n=50,017 | Odds Ratio |  |  |  | Adjusted Odds Ratio |  |  |  |
|  |  |  | OR | L | U | P | OR | L | U | P |
| ≥37w | 20 (0.08) | 37 (0.07) | 1.09 | 0.63 | 1.88 | 0.31 | 1.09 | 0.63 | 1.87 | 0.77 |
| <37w | 65 (0.26) | 88 (0.18) | 1.49 | 1.08 | 2.05 | 0.015 | 1.49 | 1.08 | 2.05 | 0.015 |
| < 32w | 40 (0.16) | 54 (0.11) | 1.49 | 0.99 | 2.25 | 0.055 | 1.49 | 0.99 | 2.25 | 0.055 |
| <28w | 24 (0.10) | 35 (0.07) | 1.38 | 0.82 | 2.32 | 0.22 | 1.38 | 0.82 | 2.32 | 0.23 |

aOR, adjusted odds ratio; L, lower limit of 95% confidence interval; U, Upper limit of 95% confidence interval; OR, odds ratio; w, weeks

**Figure 3.**
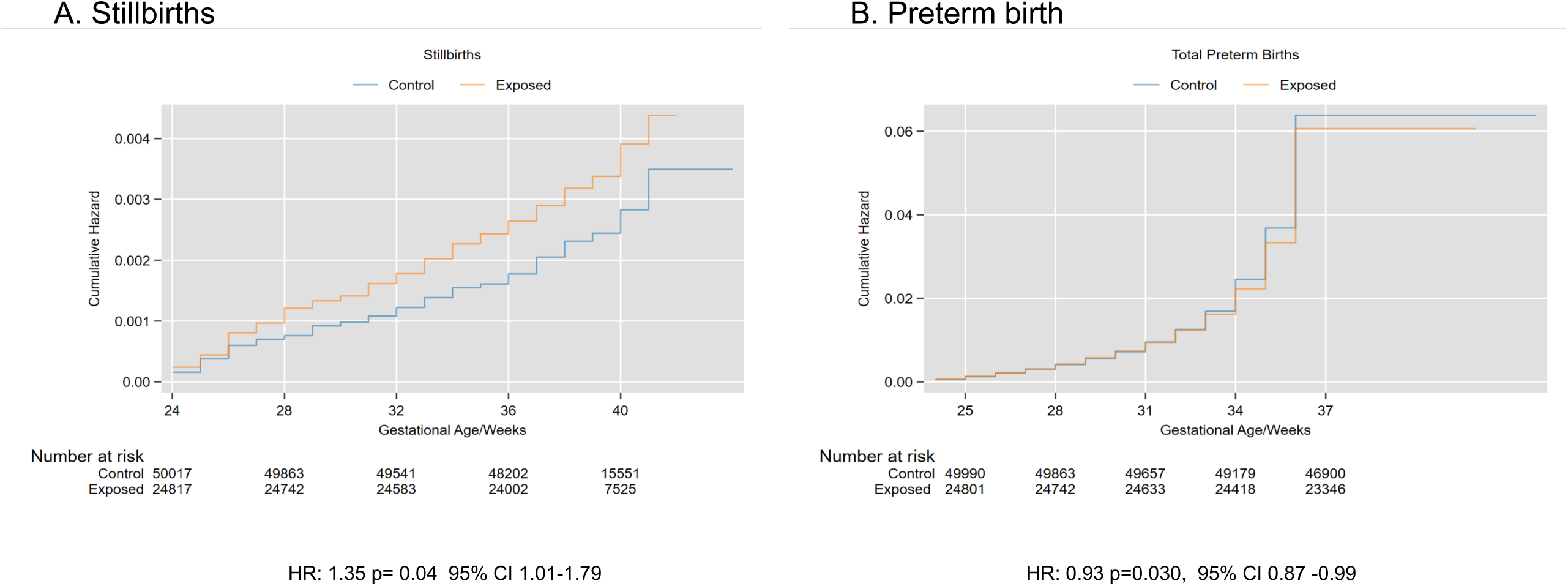
Stillbirth hazard ratio. CI, confidence interval; HR, hazard ratio

#### Total PTB < 37 weeks

There was a significant reduction in total PTB < 37 weeks for all births (5.93% vs 6.23%, aOR 0.93, 95% CI 0.87-0.99, P = 0.04; Table 2), and PTB <37w in live births (5.68% vs 6.07%, aOR 0.91, 95%CI 0.85-0.98, P = 0.009; Table 3). The hazard curve showed significant separation of the trajectories of lockdown-exposed and control groups from 33-36 weeks of gestational age (Figure 3b).

The forest plots of stillbirths and total PTB by hospital and maternity service capacity are shown in Figure 4.

**Figure 4.**
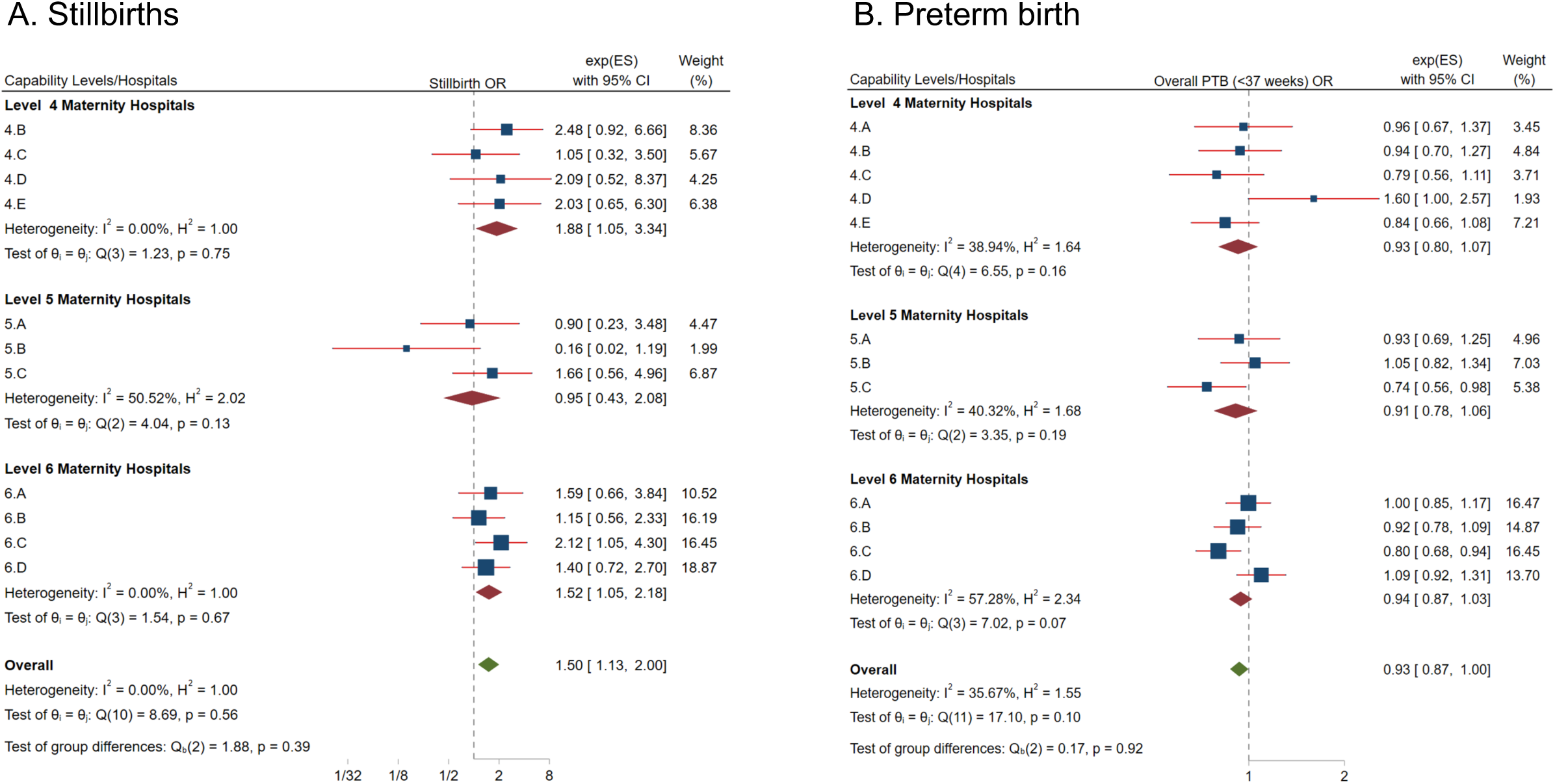
Forest plots of primary outcomes by hospital and maternity service level. 5A. Odds ratio of stillbirth 5B. Odds ratio of preterm birth < 37 weeks

### Secondary outcomes

#### PTB < 37 weeks, iatrogenic and spontaneous; all births and live births

There was no significant reduction of iatrogenic PTB < 37 weeks for all births (3.24 vs 3.42, aOR 0.92, 95 CI 0.84-1.00, P=0.08; table 2), although the reduction was significant for live births (3.01% vs 3.27%, aOR 0.89, 95% CI 0.81-0.98, P= 0.015; table 3). There was no significant difference between the exposed and control groups in rates of spontaneous preterm birth < 37 weeks’ gestation for live infants (2.68% vs 2.80%, aOR 0.94, 95%CI 0.86-1.04, P=0.26) or all infants (2.69% vs 2.82%, aOR 0.94, 95%CI 0.86-1.03, P=0.24). Forest plots of iatrogenic and spontaneous PTB <37weeks by hospital are provided in supplemental file 2.

#### PTB < 32 weeks, PTB <28w weeks; total, iatrogenic and spontaneous

There were notable trends towards lower PTB at < 32 and < 28 weeks for all births and live births, including total, iatrogenic and spontaneous PTB, but none of these reached statistical significance (Tables 2 and 3).

#### Iatrogenic births for fetal compromise (live births only)

There was a significant reduction in iatrogenic births for fetal compromise in the exposed group compared with the control group (16.98% vs 18.53%, OR 0.90, 95%CI 0.86-0.94, P < 0.001). This reduction was significant for gestational age groups ≥37, < 37, and < 28 weeks (Table 5).

**Table 5.** Iatrogenic live births for fetal compromise.

| Table 5 - Iatrogenic live births for fetal compromise |  |  |  |  |  |  |  |  |  |  |
| --- | --- | --- | --- | --- | --- | --- | --- | --- | --- | --- |
| Gestational age | Exposed<br>(n=24,732) | Control<br>(n=49,892) | Odds Ratio |  |  |  | Adjusted Odds Ratio |  |  |  |
|  |  |  | OR | L | U | P value | OR | L | U | P value |
| Total | 4,199 (16.98) | 9245 (18.53) | 0.90 | 0.86 | 0.94 | <0.001 | 0.90 | 0.86 | 0.94 | <0.001 |
| ≥ 37w | 3890 (15.73) | 8494 (17.02) | 0.91 | 0.87 | 0.95 | <0.001 | 0.91 | 0.88 | 0.95 | <0.001 |
| < 37w | 309 (1.25) | 751 (1.51) | 0.83 | 0.72 | 0.94 | 0.0050 | 0.79 | 0.69 | 0.91 | 0.0010 |
| < 32w | 52 (0.21) | 134 (0.27) | 0.78 | 0.57 | 1.08 | 0.13 | 0.68 | 0.47 | 0.96 | 0.030 |
| < 28w | 5 (0.02) | 32 (0.06) | 0.32 | 0.12 | 0.81 | 0.016 | 0.34 | 0.13 | 0.88 | 0.026 |

aOR, adjusted odds ratio; L, lower limit of 95% confidence interval; U, Upper limit of 95% confidence interval; OR, odds ratio; w, week

#### Fetal growth restriction

There was no significant difference between the exposed and control groups in the rate of FGR among all births (2.00% vs 2.02%, aOR 0.99, 95%CI 0.89-1.11, P=0.91) or live births (1.94 vs 1.98%, aOR 0.98, 95%CI 0.88-1.10, P=0.78). There was a higher prevalence of FGR among preterm infants in the exposed cohort, but this did not reach statistical significance for any of the gestational age subgroups. (See Supplemental file 3).

#### Apgars < 7 at five minutes and admissions to SCN and NICU (live births)

There was no significant difference in low Apgar scores between the live infants in the exposed and control groups. There was a significant reduction in admissions to the SCN in the exposed group (11.53% vs 12.51%, aOR 0.91, 95%CI 0.87-0.96, P< 0.0001). There was no significant difference in admissions to the NICU.

#### Pregnancy care indicators (all births)

A significant proportion of women in the exposed group had their first antenatal visit ≤ 12 weeks gestation (74.44% vs 58.96%, aOR 2.05, 95%CI 1.98-2.12, p< 0.0001). There was a significant increase in infants born before arrival to hospital, from 0.50% to 0.57% (aOR 1.25, 95% 1.01-1.54, P= 0.046) in the lockdown exposed group.

### Run chart analysis

There was a median of 801 total births per week during the run chart analysis period. There was a significant downward shift in new pregnancies during March-April 2020 at the onset of lockdown, followed by a significant rebound above the median in May and June (Figure 5a). Run charts of primary outcomes by week of cLMP confirmed the temporal relationship between lockdown restrictions and the significant changes observed in the cohort analysis. There was a significant shift to more stillbirths after the onset of lockdown (Figure 5b), and shifts to fewer preterm births <37 weeks (Figure 6a). Run charts of the secondary outcomes were consistent with the cohort analysis, except for the outcome measure of first antenatal visit ≤ 12 weeks (Figure 7). The run chart displaying rates of first antenatal visit ≤ 12 weeks demonstrated an upward secular trend that predated the lockdown (Figure 8a).

**Figure 5.**
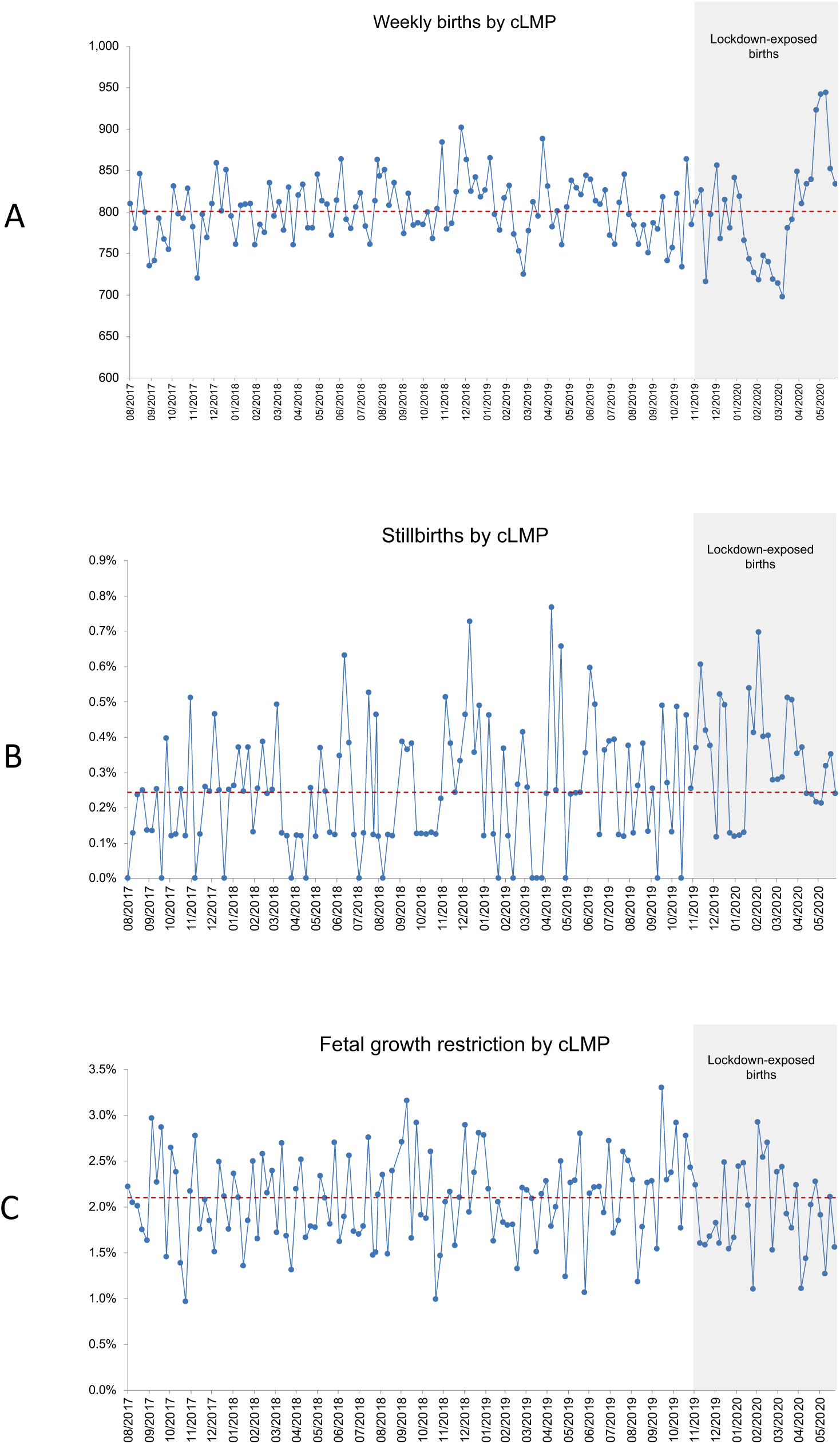
Run charts of weekly births, stillbirths and fetal growth restriction by cLMP. 5A. Weekly births by cLMP 5B. Stillbirths by cLMP 5C. Fetal growth restriction by cLMP cLMP, calculated last menstrual period

**Figure 6.**
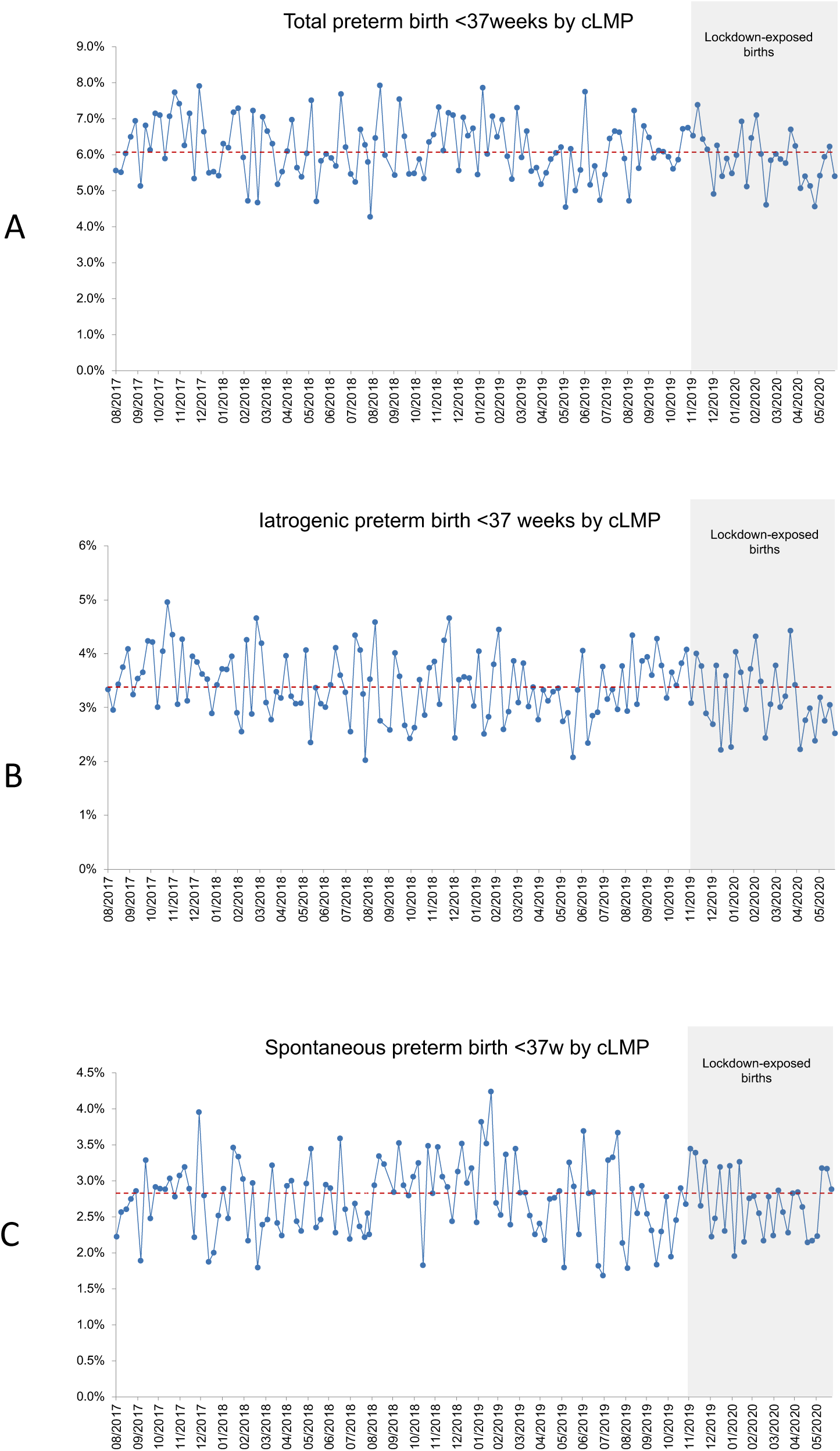
Run charts of preterm births < 37 weeks by cLMP. 6A. Total preterm birth by cLMP 6B. Iatrogenic preterm birth by cLMP 6C. Spontaneous preterm birth by cLMP cLMP, calculated last menstrual period

**Figure 7.**
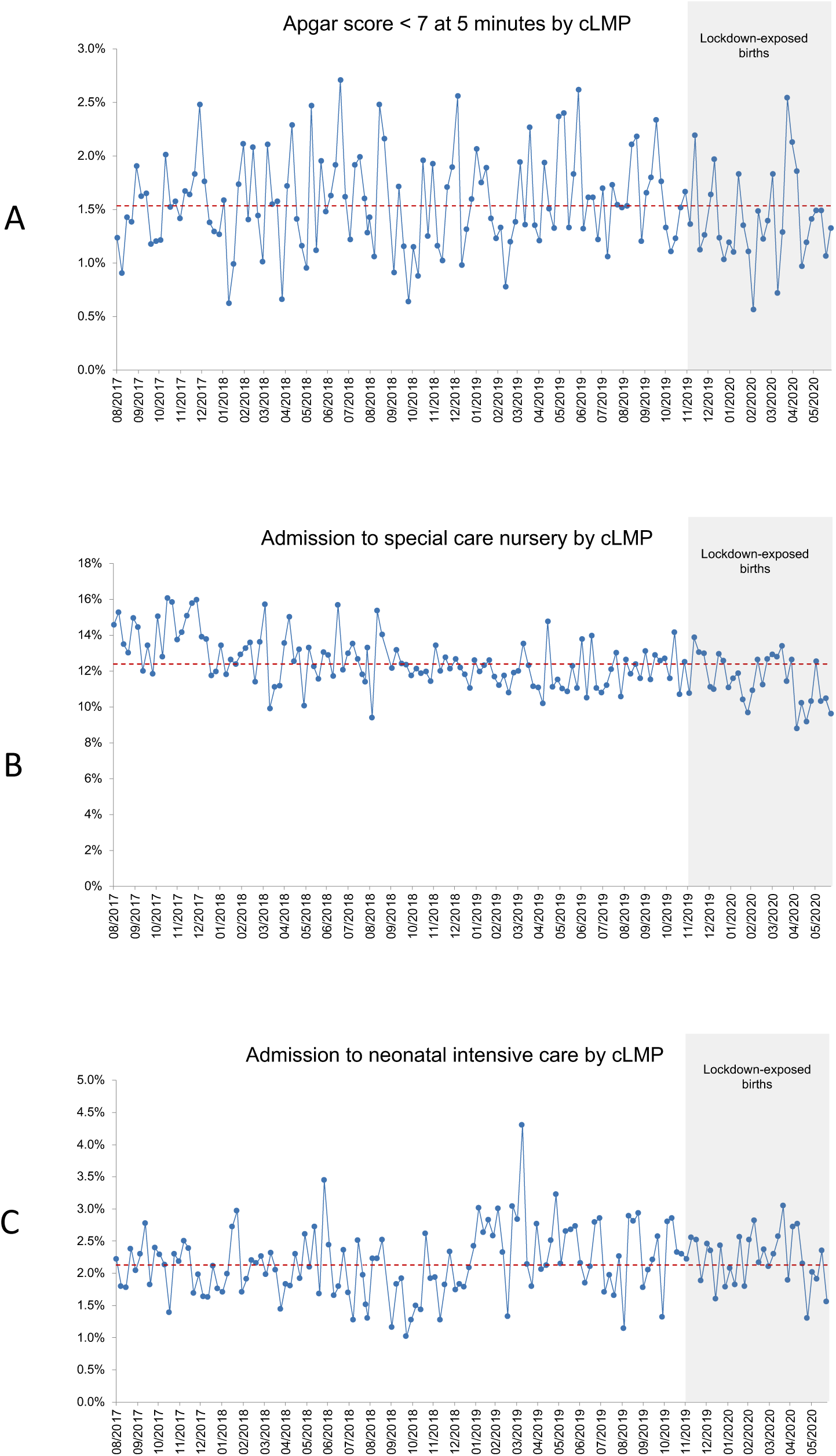
Run charts of newborn outcomes by cLMP. 7A. Apgar score < 7 at 5 minutes 7B. Admission to special care nursery 7C. Admission to neonatal intensive care cLMP, calculated last menstrual period

**Figure 8.**
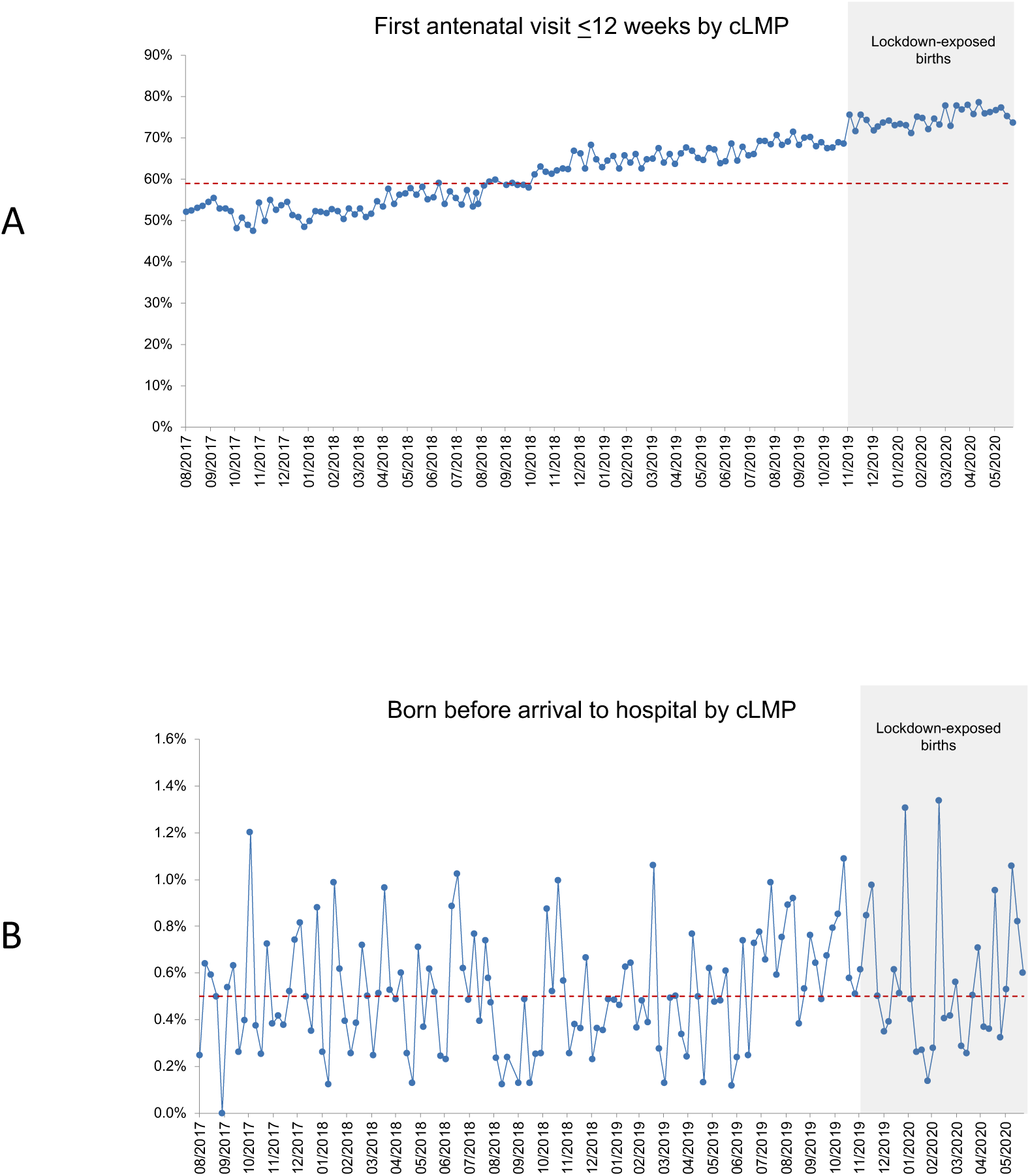
Run charts of pregnancy care indicators by cLMP. A. First antenatal visit ≤12 weeks gestation by cLMP B. Born before arrival by cLMP cLMP, calculated last menstrual period

## Discussion

Our multicentre cohort study assessing the impact of lockdown restrictions and altered antenatal care in the absence of high COVID-19 disease rates, demonstrated a significant increase in preterm stillbirths among women who were pregnant during the 2020 lockdown. This was accompanied by a significant reduction in total PTB < 37 weeks, including a significant reduction in iatrogenic PTB < 37 weeks for fetal compromise. FGR has the largest population attributable risk for stillbirth, with a fivefold higher stillbirth risk if FGR is undetected antenatally.^24^ The overall prevalence of FGR across all births was no different between the exposed and control groups, nor was there any significant difference in the rate of intrapartum stillbirths. It is plausible that fewer episodes of in-person care during lockdown reduced the detection of growth restricted fetuses via routine obstetric examination or maternal report of decreased fetal movements.^25^ Taken together, these results suggest that the increase in stillbirth may be due to a failure of detection, appropriate surveillance and timely delivery for preterm FGR infants during the lockdown period, rather than a rise in the prevalence of FGR, or deficiencies in intrapartum care.

This rise in stillbirth comes despite significantly higher rates of early engagement in antenatal care in the lockdown group. The higher rate of first antenatal visits by 12 weeks’ gestation during the lockdown period may be due to greater access to appointments facilitated by telehealth or may be a continuation of a long-term trend unrelated to the pandemic. A single health service study from Melbourne examining maternal and perinatal outcomes after the introduction of maternity telehealth concluded that there were no adverse impacts of remote consultations, though this was not adequately powered for rare outcomes such as stillbirth.^26^ Our analysis demonstrate consistent trends to higher stillbirth rates and lower preterm birth rates across the majority of individual hospitals. The increase in stillbirth might be due to a maternal reluctance to present to hospital for decreased fetal movements or other medical or obstetrics concerns, rather than institution-specific barriers to in-person care. The significant increase in babies born before arrival to hospital during lockdown suggests that women were delaying presentation in labour - possibly due to fears about the risks of inpatient care, concerns regarding restrictive visiting policies for support persons in maternity wards, or home care responsibilities for other children. Not surprisingly, increases in stress and anxiety across the entire maternity sector during 2020 has been reported.^27^ Further research is needed to understand whether the increase in preterm stillbirths is related to health service access, utilization, altered maternal pathophysiology, or other social and environmental factors.

We found a nonsignificant reduction in spontaneous preterm birth of a magnitude similar to that reported in a meta analysis.^1^ This contrasts with findings from two institution-based studies from the United States that did find a significant reduction in spontaneous PTB.^11,28^ However, these studies had different baseline rates of PTB and altered demographic profiles, and higher COVID-19 caseloads). An earlier local report from a single health service in Melbourne also reported a reduction in preterm birth, without a significant increase in stillbirth.^8^ This study differed in inclusion criteria (included multiple pregnancies and births < 24 weeks), as well as sample size, study period, case mix, and geographical coverage. Our present Melbourne-wide dataset was sufficiently powered to examine stillbirth as a primary outcome, overcoming many of the limitations of single institution studies. The forest plots demonstrated consistent trends across each hospital and service capability levels, demonstrating that the changes in primary outcomes are most plausibly due to the effect of lockdown rather than any other variable.

An incidental finding was a marked decline in weekly pregnancies at the onset of the lockdown in March and April 2020, followed by a rebound in May and June 2020. It appears that many women who might have otherwise planned to conceive when the national state of emergency was announced on 16 March 2020 delayed conception until June, when the first lockdown restrictions eased. This shift in conceptions would also explain the small increase in median maternal age of one month in the lockdown cohort. This phenomenon of a steep decline in births followed by a “baby boom” is a well-established pattern associated with pandemics.^29^

### Strengths

Melbourne is a unique case study for the impact of pandemic restrictions on pregnancy outcomes due to the stringency and length of lockdown and the lack of significant health system strain or COVID-19 maternity caseload. Our adverse results can therefore be solely attributed to indirect effects of the pandemic. Our report has the strengths of a large sample size and complete ascertainment of all public hospital births. In any study examining preterm birth during the pandemic, it is critical to have geographical coverage of all levels of maternity services as patient presentation and interhospital transfer patterns may be altered. Our maternity services are linked via a statewide perinatal retrieval service that facilitates the rapid transfer of women with high-risk pregnancies to appropriate level 6 facilities. This allows us to assume capture of all births < 31 weeks’ as private and regional hospitals typically do not have capacity to care for very preterm infants.

Our analysis also addresses the hidden perinatal mortality inherent in studies that report reductions in live preterm births without including stillbirths.^30^ We heeded the recent call of the International Stillbirth Alliance to report both PTB and stillbirths, distinguish antepartum from intrapartum stillbirths, and achieve an adequately sized control group.^31^ Furthermore, our study defined cohorts by cLMP rather than date of birth to allow proper calculation of outcome rates using women with similar timing of exposure to lockdown conditions, and to avoid skew in ascertainment of preterm or term births at the beginning and end of the birth data collection periods respectively.

### Limitations

The major limitation of our study is the omission of exclusively private hospital births, which make up about one quarter of total births in Melbourne. Private hospitals care for low to moderate risk pregnancies but have markedly different practices compared with the public sector, including higher rates of term inductions of labour and Caesarean births and higher rates of undetected FGR.^32^ Our results are therefore not generalizable to perinatal outcomes in private hospitals.

## Conclusions

Lockdown restrictions have significant impacts on perinatal outcomes, independent of the effect of COVID-19 disease. Our study shows that pregnant women exposed to lockdown in a high income setting were less likely to have an iatrogenic preterm birth for suspected fetal compromise but more likely to have a preterm stillbirth. Health services, consumers, practitioners, and health policy makers must consider the consequences of our pandemic response on maternity care as we continue to navigate the second year of the COVID-19 pandemic.

## Data Availability

Non-identifiable individual participant data is available on request from the Austin Health Human Research Ethics Committee and the Mercy Health Human Research Ethics Committee. The study investigators may contribute aggregate and non-identifiable individual patient data to national and international collaborations whose proposed use of the data has been ethically reviewed and approved by an independent committee and following signing of an appropriate research collaboration agreement with the University of Melbourne.

## Contributors

LH, SW, JS, CW, DR, BM, KP and SP conceived of the original collaboration, LH designed, coordinated, and acquired funding for the study; all authors contributed to the data analysis plan; LH, SP, JS, DR, CW, PS, JF, MM collected the primary data; MM, NP, SP, LH performed the data cleaning and coding; LH and MM performed the data analysis; SW, NP, DR, and BM verified the data analysis; MM and LH created the tables and figures; LH wrote the primary manuscript; all authors reviewed and edited the draft manuscript and approved the final submitted manuscript.

## Acknowledgements

The health services and individual hospitals contributing to the Collaborative Maternity and Newborn Dashboard for COVID-19 pandemic are:

- Mercy Health (Mercy Hospital for Women, Werribee Mercy Hospital)
- The Royal Women’s Hospital, The Women’s at Sandringham
- Monash Health (Monash Medical Centre, Casey Hospital, Dandenong Hospital)
- Northern Health (The Northern Hospital)
- Western Health (Joan Kirner Women’s and Children’s, Sunshine Hospital)
- Eastern Health (Box Hill Hospital, The Angliss Hospital)
- Peninsula Health (Frankston Hospital)

We thank the health service data managers and research midwives (Tania Fletcher, Lynn Rigg, Michelle Knight, Eleanor Johnson, Abby Monaghan, Pauline Hamilton, Roshanee Perera) for their assistance with primary data collection and Mr Andrew Goldsachs for assistance with coding of the indications for iatrogenic births.

## Supplementary files

Supplemental File 1. Bivariate analyses of covariate and perinatal outcomes

Supplementary File 2. Table of fetal growth restriction rates by gestational age

Supplemental File 3. Maternal characteristics of exposed and control stillbirth group

Supplemental File 4. Rate of fetal growth restriction among stillbirths by gestational age

Supplemental File 5. Forest plot of iatrogenic and spontaneous preterm birth < 37 weeks by hospital

