## Supplementary figures and images for "Increase in preterm stillbirths and reduction in iatrogenic preterm births for fetal compromise: a multi-centre cohort study of COVID-19 lockdown effects in Melbourne, Australia"

### Supplemental figure 1

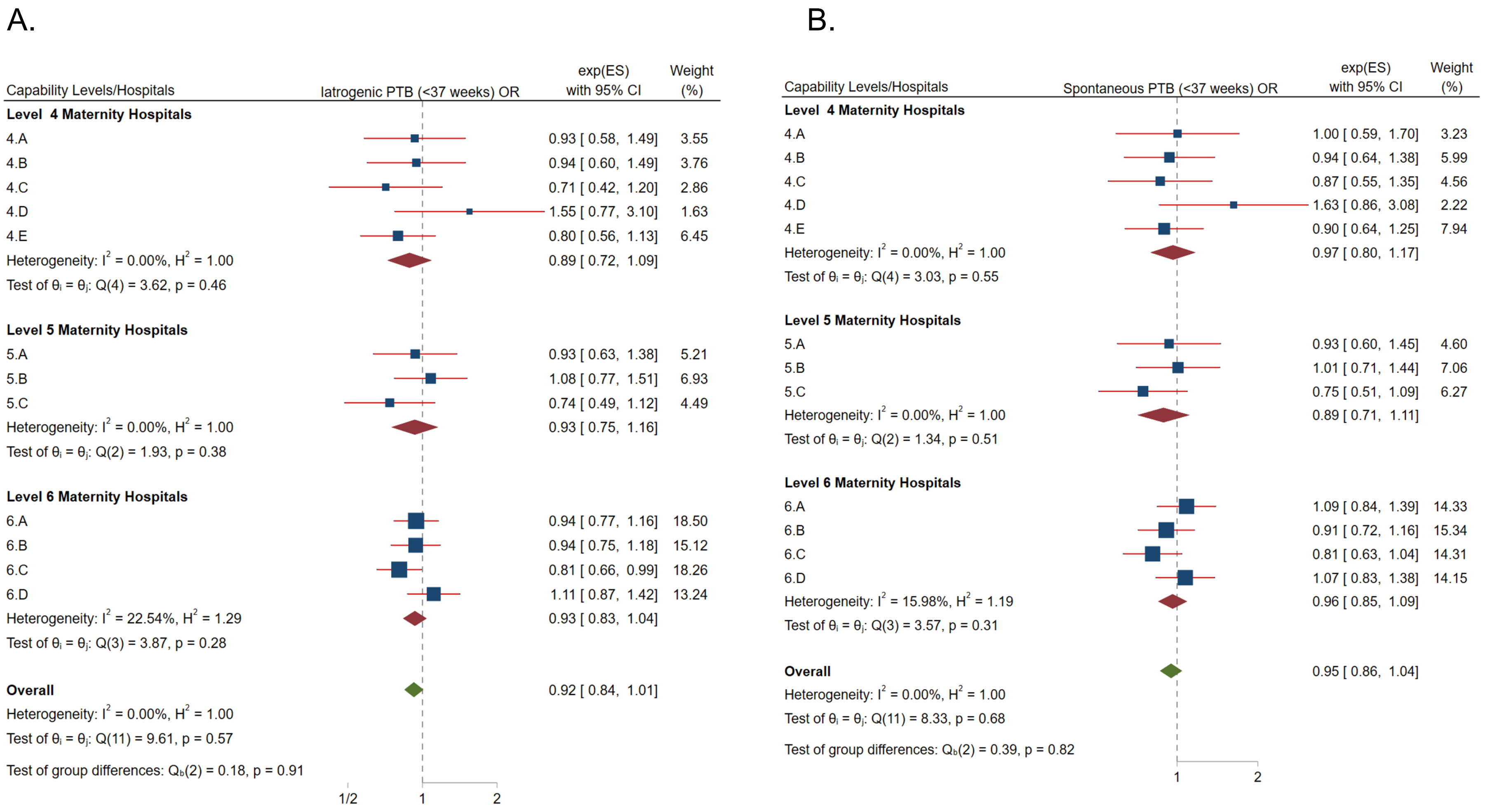
