## Supplemental tables 1-4 for "Increase in preterm stillbirths and reduction in iatrogenic preterm births for fetal compromise: a multi-centre cohort study of COVID-19 lockdown effects in Melbourne, Australia"

| **Supplemental Table 1. Bivariate Analyses of Covariate and Perinatal Outcomes** | | | | | | | | | | | | | | | | | | | | |
| --- | --- | --- | --- | --- | --- | --- | --- | --- | --- | --- | --- | --- | --- | --- | --- | --- | --- | --- | --- | --- |
|  | Stillbirths | | | | SCN/NICU admission | | | | Preterm Birth <28 weeks | | | | Preterm Birth (<32 weeks | | | | Preterm Birth <37 weeks | | | |
|  | OR | L | U | P | OR | L | U | P | OR | L | U | P | OR | L | U | P | OR | L | U | P |
| **Age Group** |  |  |  |  |  |  |  |  |  |  |  |  |  |  |  |  |  |  |  |  |
| <25 | 1.04 | 0.66 | 1.65 | 0.85 | 1.14 | 1.06 | 1.22 | <0.001 | 1.21 | 0.79 | 1.86 | 0.38 | 1.19 | 0.92 | 1.54 | 0.19 | 1.08 | 0.97 | 1.20 | 0.17 |
| 25-29 | ref | - | - | - | ref | - | - | - | ref | - | - | - | ref | - | - | - | ref | - | - | - |
| 30-34 | 0.82 | 0.57 | 1.17 | 0.27 | 0.93 | 0.88 | 0.98 | <0.01 | 0.80 | 0.56 | 1.13 | 0.21 | 0.97 | 0.79 | 1.19 | 0.75 | 0.98 | 0.91 | 1.06 | 0.66 |
| 35-39 | 0.89 | 0.59 | 1.34 | 0.58 | 0.95 | 0.89 | 1.01 | 0.12 | 1.06 | 0.72 | 1.55 | 0.78 | 1.14 | 0.91 | 1.42 | 0.26 | 1.16 | 1.06 | 1.27 | <0.01 |
| 40+ | 1.38 | 0.76 | 2.50 | 0.28 | 1.13 | 1.02 | 1.25 | <0.05 | 1.43 | 0.80 | 2.53 | 0.23 | 1.72 | 1.25 | 2.37 | <0.01 | 1.63 | 1.42 | 1.86 | <0.001 |
| **SEIFA** |  |  |  |  |  |  |  |  |  |  |  |  |  |  |  |  |  |  |  |  |
| 1 - Lowest | 1.29 | 0.83 | 2.00 | 0.26 | 1.11 | 1.03 | 1.18 | <0.01 | 0.91 | 0.61 | 1.34 | 0.62 | 0.92 | 0.73 | 1.16 | 0.48 | 1.06 | 0.96 | 1.17 | 0.25 |
| 2 | ref | - | - | - | ref | - | - | - | ref | - | - | - | ref | - | - | - | ref | - | - | - |
| 3 | 1.09 | 0.70 | 1.71 | 0.70 | 1.06 | 0.99 | 1.13 | 0.09 | 0.83 | 0.56 | 1.22 | 0.34 | 0.83 | 0.66 | 1.04 | 0.11 | 1.01 | 0.91 | 1.11 | 0.89 |
| 4 | 0.86 | 0.53 | 1.38 | 0.52 | 0.93 | 0.87 | 1.00 | 0.05 | 0.54 | 0.35 | 0.84 | <0.01 | 0.70 | 0.55 | 0.89 | <0.01 | 0.87 | 0.78 | 0.96 | <0.01 |
| 5-Advantaged | 0.83 | 0.50 | 1.38 | 0.47 | 0.80 | 0.74 | 0.86 | <0.001 | 0.56 | 0.35 | 0.90 | <0.05 | 0.66 | 0.51 | 0.86 | <0.01 | 0.80 | 0.72 | 0.89 | <0.01 |
| **Parity** |  |  |  |  |  |  |  |  |  |  |  |  |  |  |  |  |  |  |  |  |
| 0 | 1.16 | 0.85 | 1.57 | 0.35 | 1.62 | 1.54 | 1.69 | <0.001 | 1.44 | 1.06 | 1.97 | <0.05 | 1.42 | 1.19 | 1.69 | <0.001 | 1.25 | 1.17 | 1.35 | <0.001 |
| 1 | ref | - | - | - | ref | - | - | - | ref | - | - | - | ref | - | - | - | ref | - | - | - |
| 2 | 0.87 | 0.54 | 1.41 | 0.58 | 1.19 | 1.11 | 1.27 | <0.001 | 1.02 | 0.63 | 1.64 | 0.95 | 1.14 | 0.88 | 1.47 | 0.33 | 1.24 | 1.12 | 1.36 | <0.001 |
| >=3 | 1.37 | 0.82 | 2.27 | 0.23 | 1.53 | 1.41 | 1.66 | <0.001 | 2.56 | 1.67 | 3.92 | <0.001 | 2.02 | 1.55 | 2.62 | <0.001 | 1.66 | 1.49 | 1.85 | <0.001 |
| **BMI** |  |  |  |  |  |  |  |  |  |  |  |  |  |  |  |  |  |  |  |  |
| <18 | 0.53 | 0.07 | 3.81 | 0.53 | 1.28 | 1.07 | 1.53 | <0.01 | 1.48 | 0.47 | 4.71 | 0.51 | 1.37 | 0.70 | 2.67 | 0.36 | 1.43 | 1.12 | 1.83 | <0.01 |
| 18-24 | ref | - | - | - | ref | - | - | - | ref | - | - | - | ref | - | - | - | ref | - | - | - |
| 25-29 | 1.76 | 1.27 | 2.44 | <0.01 | 1.18 | 1.12 | 1.24 | <0.001 | 1.33 | 0.94 | 1.87 | 0.11 | 1.28 | 1.06 | 1.55 | <0.05 | 1.15 | 1.07 | 1.24 | <0.001 |
| 30-34 | 1.74 | 1.14 | 2.66 | <0.05 | 1.35 | 1.26 | 1.43 | <0.001 | 1.52 | 0.99 | 2.33 | 0.06 | 1.45 | 1.14 | 1.85 | <0.01 | 1.27 | 1.15 | 1.39 | <0.001 |
| 35-39 | 1.80 | 1.02 | 3.20 | <0.05 | 1.67 | 1.53 | 1.81 | <0.001 | 2.41 | 1.47 | 3.95 | <0.001 | 2.04 | 1.52 | 2.74 | <0.001 | 1.55 | 1.37 | 1.75 | <0.001 |
| >=40 | 1.41 | 0.65 | 3.06 | 0.39 | 1.93 | 1.74 | 2.13 | <0.001 | 2.06 | 1.10 | 3.89 | <0.05 | 1.56 | 1.05 | 2.33 | <0.05 | 1.51 | 1.30 | 1.76 | <0.001 |
| **Region of Birth/COB** |  |  |  |  |  |  |  |  |  |  |  |  |  |  |  |  |  |  |  |  |
| Americas | 1.02 | 0.32 | 3.22 | 0.98 | 0.58 | 0.47 | 0.70 | <0.001 | 0.26 | 0.04 | 1.89 | 0.18 | 0.55 | 0.26 | 1.16 | 0.12 | 0.60 | 0.44 | 0.80 | <0.01 |
| Australia and Associated Territories | ref | - | - | - | ref | - | - | - | ref |  | - | - | ref | - | - | - | ref | - | - | - |
| North Africa and Middle East | 1.67 | 0.98 | 2.83 | 0.06 | 0.76 | 0.69 | 0.84 | <0.001 | 1.22 | 0.71 | 2.08 | 0.48 | 0.84 | 0.60 | 1.19 | 0.34 | 0.90 | 0.78 | 1.04 | 0.15 |
| North East Asia | 0.59 | 0.24 | 1.46 | 0.26 | 0.61 | 0.54 | 0.68 | <0.001 | 0.65 | 0.30 | 1.39 | 0.26 | 0.60 | 0.39 | 0.93 | <0.05 | 0.68 | 0.57 | 0.81 | <0.001 |
| North West Europe | 0.53 | 0.17 | 1.67 | 0.28 | 0.75 | 0.65 | 0.85 | <0.001 | 0.14 | 0.02 | 0.98 | 0.05 | 0.37 | 0.19 | 0.71 | <0.01 | 0.68 | 0.55 | 0.83 | <0.001 |
| Oceania including Antartica | 1.22 | 0.61 | 2.41 | 0.58 | 1.03 | 0.93 | 1.14 | 0.56 | 1.26 | 0.70 | 2.29 | 0.44 | 0.94 | 0.65 | 1.36 | 0.75 | 0.94 | 0.80 | 1.10 | 0.44 |
| South East Asia | 1.02 | 0.61 | 1.71 | 0.95 | 0.79 | 0.73 | 0.86 | <0.001 | 0.65 | 0.37 | 1.13 | 0.13 | 0.58 | 0.42 | 0.80 | <0.01 | 0.87 | 0.78 | 0.98 | <0.05 |
| Southern and Central Asia | 1.31 | 0.92 | 1.85 | 0.13 | 1.00 | 0.94 | 1.05 | 0.89 | 0.85 | 0.59 | 1.21 | 0.36 | 0.66 | 0.53 | 0.82 | <0.001 | 0.92 | 0.85 | 1.00 | <0.05 |
| Southern and Eastern Europe | 0.50 | 0.12 | 2.03 | 0.33 | 0.69 | 0.59 | 0.81 | <0.001 | 0.78 | 0.29 | 2.11 | 0.62 | 0.75 | 0.43 | 1.31 | 0.32 | 0.80 | 0.64 | 1.01 | 0.06 |
| Sub-Saharan Africa | 1.35 | 0.65 | 2.78 | 0.42 | 0.87 | 0.77 | 0.99 | 0.03 | 1.05 | 0.51 | 2.15 | 0.89 | 0.98 | 0.65 | 1.47 | 0.92 | 0.80 | 0.67 | 0.97 | <0.05 |
| **Smoking Status** |  |  |  |  |  |  |  |  |  |  |  |  |  |  |  |  |  |  |  |  |
| Yes | 1.43 | 0.86 | 2.38 | 0.17 | 1.96 | 1.82 | 2.12 | <0.001 | 2.03 | 1.33 | 3.10 | <0.01 | 1.94 | 1.51 | 2.48 | <0.001 | 1.94 | 1.75 | 2.16 | <0.001 |
| **Interpreter Required** |  |  |  |  |  |  |  |  |  |  |  |  |  |  |  |  |  |  |  |  |
| Yes | 1.30 | 0.79 | 2.14 | 0.30 | 0.86 | 0.78 | 0.94 | <0.01 | 0.75 | 0.41 | 1.37 | 0.34 | 0.51 | 0.34 | 0.77 | <0.01 | 0.84 | 0.74 | 0.96 | <0.05 |
| **Infant Sex** |  |  |  |  |  |  |  |  |  |  |  |  |  |  |  |  |  |  |  |  |
| Male | 1.22 | 0.93 | 1.60 | 0.17 | 1.23 | 1.18 | 1.28 | <0.001 | 1.20 | 0.93 | 1.56 | 0.17 | 1.22 | 1.05 | 1.41 | <0.01 | 1.20 | 1.13 | 1.27 | <0.001 |
| Female | ref | - | - | - |  |  |  | - | ref | - | - | - | ref | - | - | - | ref | - | - | - |
| **Fetal growth restriction** |  |  |  |  |  |  |  |  |  |  |  |  |  |  |  |  |  |  |  |  |
| Yes | 10.71 | 7.48 | 15.34 | <0.001 | 4.766205 | 4.289417 | 5.295991 | <0.001 | 6.36 | 4.21 | 9.60 | <0.001 | 3.05 | 2.21 | 4.20 | <0.001 | 2.10 | 1.79 | 2.46 | <0.001 |
| outcomes includes all births (live births and stillbirths) except for admission to SCN NICU (live births only) | | | | | | | |  |  |  |  |  |  |  |  |  |  |  |  |  |

| **Supplemental table 2 - Fetal growth restriction by gestational age at birth** | | | | | | |
| --- | --- | --- | --- | --- | --- | --- |
|  | Exposed | Control | Odds Ratio | | | |
|  |  |  | OR | L | U | P |
| **Fetal growth restriction ≥ 37w** |  |  |  |  |  |  |
| All births | 426 (1.73) | 899 (1.80) | 0.96 | 0.85 | 1.08 | 0.49 |
| Live births only | 425 (1.73) | 895 (1.79) | 0.96 | 0.86 | 1.08 | 0.52 |
| **Fetal growth restriction <37w** |  |  |  |  |  |  |
| All births | 67 (0.27) | 111 (0.22) | 1.22 | 0.90 | 1.66 | 0.19 |
| Live births only | 53 (0.22) | 93 (0.19) | 1.16 | 0.82 | 1.62 | 0.40 |
| **Fetal growth restriction <32w** |  |  |  |  |  |  |
| All births | 15 (0.06) | 26 (0.05) | 1.17 | 0.62 | 2.21 | 0.63 |
| Live births only | 5 (0.02) | 16 (0.03) | 0.63 | 0.23 | 1.73 | 0.37 |
| **Fetal growth restriction <28w** |  |  |  |  |  |  |
| All births | 10 (0.04) | 16 (0.03) | 1.27 | 0.57 | 2.79 | 0.56 |
| Live births only | 3 (0.01) | 7 (0.01) | 0.87 | 0.22 | 3.36 | 0.84 |

| **Supplementary Table 3 - Maternal characteristics among control and exposed stillbirth groups** | | | |
| --- | --- | --- | --- |
| **Maternal characteristics** | **Exposed** | **Control** |  |
|  | **(n=85)** | **(n=125)** | **P** |
| Age at conception in Years, mean (SD) | 31.59 (5.39) | 31.71 (5.26) | 0.88 |
| Weight in Kg, mean (SD) | 71.86 (17.73) | 74.17(17.87) | 0.38 |
| Height in cm, mean (SD) | 162.78 (7.53) | 163.1 (7.37) | 0.77 |
| Smoking in Pregnancy, n (%) | 5 (5.88) | 11 (8.80) | 0.43 |
| **BMI Categories, n** (%) |  |  |  |
| <18 | 1 (1.28) | 0 (0) | 0.72 |
| 18-24 | 31 (39.74) | 39 (33.05) |  |
| 25-29 | 28 (35.90) | 45 (38.14) |  |
| 30-34 | 11 (14.10) | 20 (16.95) |  |
| 35-39 | 5 (6.41) | 9 (7.63) |  |
| >=40 | 2 (2.56) | 5 (4.24) |  |
| **Region of Birth, n** (%) |  |  |  |
| Americas | 1 (1.20) | 2 (1.63) | 0.82 |
| Australia and Associated Territories | 41 (49.40) | 54 (43.90) |  |
| North Africa and Middle East | 7 (8.43) | 9 (7.32) |  |
| North East Asia | 3 (3.61) | 2 (1.63) |  |
| North West Europe | 2 (2.41) | 1 (0.81) |  |
| Oceania including Antarctica | 3 (3.61) | 6 (4.88) |  |
| South East Asia | 6 (7.23) | 11 (8.94) |  |
| Southern and Central Asia | 16 (19.28) | 32 (26.02) |  |
| Southern and Eastern Europe | 0 (0) | 2 (1.63) |  |
| Sub-Saharan Africa | 4 (4.82) | 4 (3.25) |  |
| **Parity, n** (%) |  |  |  |
| 0 | 42 (49.41) | 57 (45.60) | 0.96 |
| 1 | 23 (27.06) | 47 (37.60) |  |
| 2 | 8 (9.41) | 14 (11.20) |  |
| >=3 | 12 (14.12) | 7 (5.60) |  |
| **SEIFA quintile, n** (%) |  |  |  |
| 1 - Most disadvantaged | 23 (27.06) | 35 (28) | 0.91 |
| 2 | 13 (15.29) | 17 (13.6) |  |
| 3 | 20 (23.53) | 34 (27.20) |  |
| 4 | 18 (21.18) | 21 (16.80) |  |
| 5 - Most advantaged | 11 (12.94) | 18 (14.40) |  |
| **Birthing location** |  |  |  |
| Level 4 maternity service | 23 (27.06) | 28 (22.40) |  |
| Level 5 maternity service | 26 (11.76) | 28 (22.40) | 0.6 |
| Level 6 maternity service | 52 (61.18) | 68 (54.40) |  |

| **Supplementary table 4. Rate of fetal growth restriction among stillbirths by gestational age** | | | | | | | | | | |
| --- | --- | --- | --- | --- | --- | --- | --- | --- | --- | --- |
| Gestational age | Exposed | Control | Odds Ratio | | | | Adjusted Odds Ratio | | | |
|  |  |  | OR | L | U | P | OR | L | U | P |
| Total | 15 (17.6) | 22 (17.6) | 1.07 | 0.54 | 2.14 | 0.84 | 1.11 | 0.52 | 2.35 | 0.79 |
| Term ≥37 | 1 (5.0) | 4 (10.8) | 0.45 | 0.05 | 4.30 | 0.49 | 0.48 | 0.04 | 5.21 | 0.54 |
| Preterm <37 | 14 (21.5) | 18 (20.5) | 1.09 | 0.50 | 2.39 | 0.83 | 1.24 | 0.54 | 2.85 | 0.61 |
| Preterm <32 | 12 (30.0) | 15 (27.7) | 1.52 | 0.56 | 4.10 | 0.41 | 1.75 | 0.57 | 5.42 | 0.33 |
| Preterm <28 | 7 (29.1) | 9 (25.7) | 1.26 | 0.39 | 4.06 | 0.69 | 1.44 | 0.38 | 5.55 | 0.59 |
